# Experiences of moderation, moderators, and moderating by online users who engage with self-harm and suicide content

**DOI:** 10.1101/2024.02.15.24302878

**Authors:** Zoë Haime, Laura Kennedy, Lydia Grace, Lucy Biddle

## Abstract

Online mental health spaces require effective content moderation for safety. Whilst policies acknowledge the need for proactive practices and moderator support, expectations and experiences of internet users engaging with self-harm and suicide content online remain unclear. Therefore, this study aimed to explore participant accounts of moderation, moderators and moderating when engaging online with self-harm/suicide (SH/S) related content.

Participants in the DELVE study were interviewed about their experiences with SH/S content online. N=14 participants were recruited to interview at baseline, with n=8 completing the 3-month follow-up, and n=7 the 6 month follow-up. Participants were also asked to complete daily diaries of their online use between interviews. Thematic analysis, with deductive coding informed by interview questions, was used to explore perspectives on moderation, moderators and moderating from interview transcripts and diary entries.

Three key themes were identified: ‘content reporting behaviour’, exploring factors influencing decisions to report SH/S content; ‘perceptions of having content blocked’, exploring participant experiences and speculative accounts of SH/S content moderation; and ‘content moderation and moderators’, examining participant views on moderation approaches, their own experiences of moderating, and insights for future moderation improvements.

This study revealed challenges in moderating SH/S content online, and highlighted inadequacies associated with current procedures. Participants struggled to self-moderate online SH/S spaces, showing the need for proactive platform-level strategies. Additionally, whilst the lived experience of moderators was valued by participants, associated risks emphasised the need for supportive measures. Policymakers and industry leaders should prioritise transparent and consistent moderation practice.

**Author Summary:** In today’s digital world, ensuring the safety of online mental health spaces is vital. Yet, there’s still a lot we don’t understand about how people experience moderation, moderators, and moderating in self-harm and suicide online spaces. Our study set out to change that by talking to 14 individuals who engage with this content online. Through interviews and diaries, we learned more about their experiences with platform and online community moderation.

Our findings showed some important things. Firstly, individuals with declining mental health struggled to use tools that might keep them safe, like reporting content. This emphasised the need for effective moderation in online mental health spaces, to prevent harm. Secondly, unclear communication and inconsistent moderation practices lead to confusion and frustration amongst users who reported content, or had their own content moderated. Improving transparency and consistency will enhance user experiences of moderation online. Lastly, users encouraged the involvement of mental health professionals into online moderating teams, suggesting platforms and online communities should provide training and supervision from professionals to their moderation staff. These findings support our recommendations for ongoing changes to moderation procedures across online platforms.

## 1. Introduction

As online platforms facilitate connections between individuals on a global scale, it becomes crucial to implement practices that maintain safe environments through content moderation [1, 2]. Platform-level content moderation is used to regulate user-generated content across entire platforms, and can involve automated tools, algorithms, and human moderators working to apply overarching policies and standards. Alternatively, community-level content moderation involves active participation of a platforms’ users, through reporting or flagging content that they find inappropriate, or against the rules of specific spaces or communities within platforms. These moderation actions at both platform and community level may be particularly relevant in controlling engagement with sensitive, harmful or illegal material, including self-harm and suicide content and discourse around it.

### 1.1. Moderation Effectiveness

To regulate online community spaces, moderators play a vital role in screening content, identifying problematic users, and enforcing rules [3]. The mechanisms employed, such as blocking or removal of content, and semi- or permanent-banning of users are thought to ensure the availability of high-quality content, whilst limiting the presence of harmful material [4]. However, experiences and outcomes of these moderating actions can vary for online users. For example, when Facebook and Instagram introduced a total ban of graphic self-harm imagery, sentiment analysis revealed an increase in anger, anticipation, and sadness in the associated Twitter [renamed ‘X’] discourse [5].

Complexity in moderation decision-making is particularly evident on platforms or online environments focused on mental health. Moderation in these spaces must balance the responsibility of protecting users from triggering content, with the provision of space for social support and recover [6]. The effectiveness of current moderation practices, including outright content bans in mental health spaces, have been further questioned. Multiple studies have investigated how users seek out potentially harmful pro-eating disorder content online and evade moderation systems by posting content using no text, or alternative hashtags [7, 8]. This practice is also prevalent amongst self-injury related posts on Instagram, where the use of ambiguous and unrelated hashtags leads to graphic self-harm imagery without proper content warnings [9]. Additionally, research shows that even where platforms proactively search for and remove harmful content, this can be inefficient, as content reappears on the same or alternative platforms [10, 11].

### 1.2. Who Should Moderate Content?

There are also concerns regarding the allocation of responsibility for moderating online spaces. Platform-level moderation usually relies on paid humans, although these mechanisms are sometimes automated. However, within peer-driven communities, users themselves usually moderate content, often in voluntary roles. While several benefits to the moderator role have been reported, including feelings of altruism, having a sense of purpose [12] and – in the case of lived-experience moderators – receiving validation of their own experiences [13], concerns have also been raised for both community and platform level moderators’ about the impact on their own mental health [6, 12].

A recent study by Spence et al [14], found 40.8% of 213 online platform-level content moderators were exposed to distressing content daily, and that moderators showed a dose-dependent relationship between frequency of exposure to distressing content and psychological distress and secondary trauma. However, their findings also revealed, that after accounting for work factors in the analysis, including access to supportive colleagues and feedback about their work, the relationships between exposure and psychological distress and secondary trauma failed to remain significant, indicating that a supportive work environment may ameliorate negative effects.

Some platforms and sites, such as Google and Facebook, have begun to implement strategies to address the risks to their content moderators, such as training on working with sensitive content, recommended access to both individual and group counselling services, and suggested ‘wellness breaks’ [15, 16]. However, the effectiveness of these interventions for moderator wellbeing, remains unclear. Additionally, whilst large platforms are able to dedicate resources to moderator care, there are less viable options available to communities hosted on those platforms, or to smaller individual platforms.

Even within larger platforms, it may also be challenging to ensure uniform care across all moderators, due to varying work environments and individual needs. For instance, diversity in preferences of moderators was shown by Saha et al [16]. Some moderators desired support from mental health experts and welcomed the idea of having trained professionals within the moderation team. However, other moderators emphasised that these spaces should not provide medical advice to users, and that moderators with lived experience of mental health problems can create the supportive community users are looking for, without additional help. This highlights the differing needs amongst moderators in mental health spaces and emphasises the need for further understanding of individual moderator narratives to optimise their experiences.

### 1.3. The Online Safety Bill

Policy and regulatory frameworks also play a significant role in the expectations and responsibilities of online platforms in moderating content. In the United Kingdom, there has been a growing emphasis on addressing online harms, through the recently enacted *Online Safety Bill* [17]. Key points within this legislation include the need for platforms to remove self-harm and suicide content, implement measures to minimise user exposure to harmful content, uphold a duty of care towards users, and provide individuals with improved mechanisms to report harmful content.

To date, limited research has been conducted exploring experiences of moderation by users who engage with online mental health content or those who moderate it. Notably, existing findings suggest that moderation is essential to mental health online spaces [6] but that current approaches may be ineffective, and, in some cases increase vulnerability to harm for users and moderators. For instance, content posted with limited information in order to avoid automatic removal systems, increases the risk to users of unexpectantly encountering potentially triggering content [7], and moderators may experience emotional distress and burnout [13]. A noticeable research gap exists in understanding user perspectives on the moderation of SH/S related content online, particularly user experiences of having their own SH/S related content moderated (reported or removed), reporting others’ SH/S content, or seeing others’ SH/S content being removed. Understanding the first-hand perspective of users engaging in mental health online spaces is essential for informing effective, responsible, and user-centric moderation strategies in the digital space.

### 1.4. Aim of the study

This study involved thematic analysis [18] of qualitative interview transcripts to gain insights into participant experiences with moderation and moderators in the context of engaging with SH/S content online. By examining these perspectives, this research aimed to deepen our understanding of how moderation practices impact user experiences and consider how this can inform the development of industry guidance.

## 2. Results

### 2.1. Participants

Fourteen participants took part. The sample had diversity in terms of age and ethnicity, and relatively good representation of gender (Table 1). All participants completed a baseline interview, eight participants completed midpoint and seven completed endpoint interviews (see [19]). Participants returned monthly diaries. Due to participant dropout, overall 44 diaries were anticipated, with a total of 31 (70.5%) diaries actually returned. All interview transcripts and diary data were used in this analysis.

**Table 1.** Demographic characteristics of participants who took part in the DELVE study.

| Demographic Variable | Total N (%) |
| --- | --- |
| **Gender** |  |
| Female | 10 (71.4) |
| Male | 4 (28.6) |
| **Ethnicity** |  |
| Asian British | 2 (14.2) |
| Black British | 1 (7.2) |
| White British | 7 (50.0) |
| Asian Other | 2 (14.2) |
| Black Other | 0 (0.0) |
| White Other | 1 (7.2) |
| Mixed Race | 1 (7.2) |
| **Age** |  |
| 16-17 Years | 1 (7.2) |
| 18-24 Years | 7 (50.0) |
| 25-35 Years | 0 (0.0) |
| 36-45 Years | 4 (28.6) |
| 46-54 Years | 2 (14.2) |
| 55+ Years | 0 (0.0) |

### 2.2. Thematic Analysis

Results are visually depicted in Figure 1. by theme and subtheme. These are explored in more detail below.

**Fig 1:**
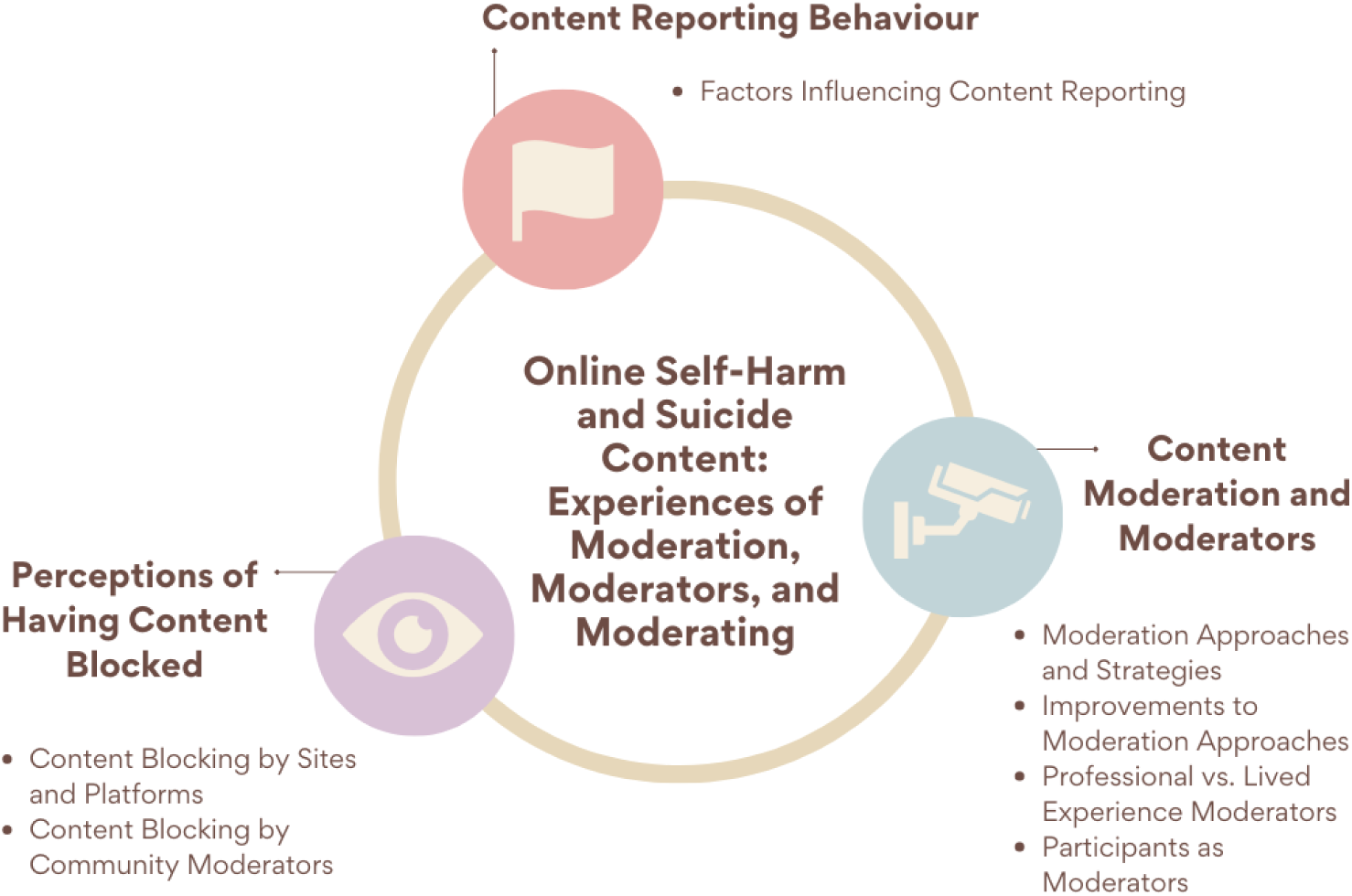
Visualisation of Themes and Subthemes.

#### 2.2.1. Content Reporting Behaviour

##### 2.2.1.1. Factors Influencing Content Reporting

Several participants described instances where they didn’t report SH/S content. One participant described how their non-reporting was influenced by a lack of awareness about how and where to report on certain platforms/sites (IDH). However, participants also shared that they did not consciously consider reporting SH/S content when online (IDC, IDH, IDM, I33), due to their poor mental state. In fact, during challenging times, users described becoming preoccupied with this material or deliberately seeking it out, thereby prioritising the perceived benefits of content over reporting it:

> ‘I think in that moment it didn’t really cross my mind [to report content about suicide methods]. I was too focused on the content itself and what it was giving.’ (IDJ)

A couple of participants described how decisions to report SH/S content were multifaceted, one stating how the choice to report would change depending upon their mental health state:

> ‘Part of me thinks they should be reported and another part of me thinks, well, freedom of speech and they should stay. So I most probably feel that they should stay, on balance. However, I think that’s one of those things that could change and fluctuate depending upon my own mental health.’ (IDC)

Participants who recalled reporting content described no hesitation in reporting general online content considered morally wrong, such as racist comments or images. However, decisions to report SH/S content were often more selective. One user (IDD) reflected on the emotional impact of content on a pro-suicide forum affecting their decision: ‘Cause like if it does affect me then… I like reduce time on it or report it to one of the moderators for example’ (IDD). Another (IDM), described how their lack of knowledge about SH/S content they came across made them refrain from reporting:

> ‘Why didn’t you report it [graphic self-harm image]?’ (Interviewer)

> ‘I didn’t know much about it [graphic self-harm image] and so I decided maybe to just not show it to anyone.’ (IDM)

Considering the implications for users posting the SH material, this participant also expressed reluctance to report images of SH without first understanding the posters’ circumstances and suggested having a conversation with them may be more appropriate than reporting:

> ‘I think you would have to talk to them, maybe see where they’re at and maybe you can report, or maybe take it to the next step with them [rather] than reporting them and then going to them. I don’t think it would be appropriate to [just report] them.’ (IDM)

Despite concern for content posters’ wellbeing, and attempts not to stigmatise their experience, there was general agreement amongst the other participants that images or videos of SH/S were largely inappropriate and justified reporting, sometimes due to their influence over participants’ own SH/S behaviours:

> ‘I can get it if someone is saying oh, I’m really suicidal, I won’t report that, because maybe they need someone to talk to, if they are posting images on that though, you can’t be doing that… And also there are lots of videos of people trying to kill themselves, they’re on the internet, that’s another thing you can spiral into that, there are sites just full of people dying…I think it was on Twitter, I ended up on a video and then it went to another video and then I ended up on a site and then yeah…’ (IDK)

> ‘…Ok and how did that make you feel sort of seeing that play out?’ (Interviewer)

> ‘I think I took an overdose afterwards’ (IDK)

One participant (IDI) described how reporting served not only as a means of addressing the issue of SH/S content posted online, but also as a way to raise awareness and educate the person responsible for the content: ‘it also, hopefully, tells that person that what they were posting [images] might have been sensitive towards someone’ (IDI).

One participant reflected on how following predetermined methods for reporting, like using checkboxes, allowed the process to be less biased, and suggested an attempt to detach themselves from the feelings of guilt associated with reporting an online peer:

> ‘What I’ll do is I’ll click all those [check] boxes and I’ll try to desensitise it or make it less personal or not take it personally. So, I try to be as objective as possible about it.’ (IDE)

When asked, those who had reported online content did describe it as a straightforward process in practical terms, with some sites having specific options for recording if the content was related to SH/S. However, one participant (IDE) acknowledged how it would be impractical to report all content that was offensive or inappropriate, and therefore they felt they had to make careful decisions about what they reported and when, although they did not detail how they made those choices, other than to note they were more likely to report users with significant follower counts: ‘Yeah, when I see something that I don’t like, like self-harm and they’ve got lots and lots of followers then I do report stuff’ (IDE).

Some of the participants who previously had not reported content, mentioned during interviews that they would now report SH/S content they came across online, using the platform guidelines to direct them (IDH, IDJ, IDC). It is unclear what motivated this change, though there was some indication it may be explained by involvement in the research itself. For instance, IDE described increased awareness of content that may be harmful, and being more conscious of its effects:

> ‘My uses have changed massively because it’s really [I’ve] been more aware, but also, I’m more conscious of it. So when you are a user you just look at it and think, ‘Okay.’ But when you do something like [taking part in this research study] you think, ‘Hang on, actually there’s another dimension to it and is there harm or should I report?’ So you become a social media – I don’t know if it’s an influencer – but you become a sort of [an] active or civic user.’ (IDE)

Some participants also described the consequences of reporting other users’ content. While a couple described content being taken down (IDK, IDI), or the user being ‘kicked off’ (IDD) a group, others commented on specific site/platform actions, stating Facebook do ‘nothing’ (IDB) when reporting nuisance pages, or that reporting on Tumblr felt ‘completely pointless…because nothing would happen’ (IDF).

IDI acknowledged that although the automated process of reporting using Instagram options made it relatively easy to do, it left no room for nuance in describing the content: ‘I don’t think there’s much of an opportunity to fully explain anything or anything like that’ (IDI). Additionally, despite reported SH/S content being removed on Instagram, users were able to continue posting without consequence:

> ‘[they can] carry on posting as normal anyway. So it just means that they can post it again’ (IDI)

#### 2.2.2. Perceptions of Having Content Blocked

##### 2.2.2.1. Content Blocking by Sites and Platforms

Several participants, (IDB, IDN, IDL, IDK, and IDG) were able to describe their experiences of having content reported online, including content expressing suicidal ideation (IDB, IDK), privately posted images of SH (IDL), and blogs about depression and self-harm (IDN). Participants described receiving differing levels of information from sites when their content had been blocked, with some feeling that the rules felt unclear:

> ‘I have like one or two blocked [blogposts], but I don’t know why. I don’t know which words [are] the restriction things. I just can’t post it out and it remains in the draft.But I think I’m just writing a normal thing, but maybe they have like a key word ban’ (IDN).

IDK, who had once posted stating they were ‘struggling’ on Twitter [X], had this reported by another user and criticised the platform’s ‘one-size-fits’ all response:

> ‘[It’s] a bit pointless to be honest, like suicide hotlines are not going to solve your issues, I think that’s the problem… but it’s [why they are struggling] about I can’t afford this, I can’t afford my house, like what am I going to do, so I found it a bit hard but thanks for sending me this, do you know what I mean? (IDK)

IDL, who used Instagram to post images privately, documenting and tracking their SH journey, recalled instances of content being blocked without warning. Removal of content interfered with their ability to monitor their well-being, and disrupted their sense of control and ownership:

> ‘…I think it’s like under [the] notification page they’ll just say, your post has been removed because of a violation. And yeah I think it’s quite frustrating because I don’tknow what I’ve written so I can forget what it was about…because I mean even if they said that, I want to remember and recall how I was feeling and the tough times I’d been through. But they don’t give you any warning at all, they just take it’ (IDL).

However, it seemed the platform was inconsistent with their moderation of these images, leaving the user uncertain what the consequences might be when posting:

> ‘…only sometimes they take it down, sometimes they don’t. It’s kind of confusing, I don’t know what they are [doing]’ (IDL).

Interestingly, one participant who had content blocked on Twitter [X] due to the use of bad language found the situation ‘quite funny to be honest’ (IDK), perhaps highlighting the difference in emotional response to content that is less personally significant.

One participant who had no personal experience of their content being reported or blocked by platforms/sites, also reflected on the potential emotional impact if it were to happen, expressing they would feel ‘disempowered’ and as if ‘you’re not being heard’ (IDC).

##### 2.2.2.2. Content Blocking by Community Moderators

In some cases, participants chose to engage with SH/S content in spaces regulated by community moderators, as well as by the sites/platforms themselves. These moderators had responsibility for enforcing community rules and guidelines, which were sometimes more specific or stricter than those set by the site/platform (e.g., individual Facebook groups on Facebook).

Two participants shared experiences of having content removed from these spaces. Both were left feeling confused and frustrated due to inconsistent and unclear moderation policies. For instance, one had (IDG) posted ‘what is the point?’ on a forum, which led to moderators removing their post and contacting them with resources of support. The participant expressed feeling ‘… just like a criminal. Well not a criminal, but like I’ve really done something wrong.’ (IDG). They also seemed surprised to see other content on the site they deemed more inappropriate hadn’t been removed.

The other participant (IDB) recalled two instances of moderators blocking content in an online private Facebook group that provided SH peer support. Initially, they posted a suicide note on the group, and moderators replaced it with a supportive post, tagging the participant. This action resulted in an influx of messages of support from other users. However, this felt intrusive, overwhelming and made the participant feel guilty, placing further strain on their mental state:

> ‘it was driving me up the wall, I couldn’t, I was like I’ve made my decision, I know what I’m doing. I don’t want you to all tell me to stop doing it, I just want some help with [suicide plans], can you please just stop because you’re just making me feel guilty and you’re not helping me, you’re not helping. Obviously it was nice but in that state I just couldn’t take it, I felt it was awful, I didn’t like it at all.’ (IDB)

In a separate event, the participant had a post removed by moderators when in disagreement with another user about them (IDB) giving mental health advice. The participant described feeling frustrated as a result of this interaction and that moderation could be biased:

> “…she’s (other user) like, “it’s so dangerous, you’re going to kill someone”, I’m like, so are you, like it says in my post I’m suicidal, is that (users’ comment to her) really helpful. And I started to get really wound up, and then the people that were managing the page took my comment off and blocked me from commenting on it anymore but allowed for her to continue which just drove me insane because I felt that was so unfair.” (IDB)

#### 2.2.3. Content Moderation and Moderators

##### 2.2.3.1. Moderation Approaches and Strategies

As participants generally perceived that some SH/S content should be accessible online, regulation and moderation of content was viewed as a necessary part of making those spaces safe:

> ‘…there is a place for it [SH/S content] online. There is a place but it needs to come from a very supportive place and it needs to be highly monitored and regulated.That’s what I’ve generally learnt. The safer places to be online and the safer places to deal with it are when it is more moderated and looked after.’ (IDB)

> ‘I don’t feel that it [online SH/S spaces] should be [taken] away because if it’s moderated correctly then it’s not doing anybody any particular harm.’ (IDD)

Participants, owing to their different encounters with SH/S content online, experienced varying interactions with moderators across platforms. Three participants (IDA, IDB and IDC) who regularly engaged with a self-harm support group hosted on Facebook, mentioned being drawn to their moderation methods, due to their clear, friendly approach, and close monitoring of content:

> ‘I tend to stick to [self-harm support Facebook group] because I know that it’s heavily moderated’ (IDB)

> ‘… it’s clear from the onset. They’re completely honest and they say this is a moderated Facebook page, so you know where you stand…A couple of days ago,somebody said about direct messaging and… very soon, a moderator came on and just said, ‘Can I just remind you?’, and he phrased it really nicely.’ (IDC).

Additionally, IDB expressed an advantage of using the self-harm support Facebook group was its rapid content moderation, which reassured them it was safe:

> ‘The moderation like I say this is why I tend to use [self-harm support Facebook group] more than anything else because… it’s so heavily moderated and monitored we don’t really find it [community argument posts]. And if anything like that does crop up on [self-harm support Facebook group] it’s gone within minutes you know. An hour is long for it to have still been on there.’ – IDB

However, it is important to note that these perspectives may have been subject to bias within the sample. Notably, IDA and IDB who regularly engaged with the self-harm support Facebook group, also moderated for them in a voluntary capacity, which may have influenced their views on effective approaches.

Participants also provided insights into the moderation practices of prominent platforms such as Twitter [X] and Facebook. Interestingly, participant experiences seemed to contradict one another. For instance, one participant criticised Facebook for using ‘a carpet ban’ (IDC) when it came to SH/S content. In contrast, another participant reported that moderation practices on Facebook were relaxed, though it is unclear whether participants were referring to community or platform moderation in these circumstances:

> ‘On Twitter [X], if you’ve reported… like graphic images or anything like that, it will be blocked, but TikTok and Facebook they’re very lenient about stuff.’ (IDK).

##### 2.2.3.2. Improvements to Moderation Approaches

Participants identified areas of moderation that they would like to see improved across platforms. One common concern for participants was the lack of transparency and consistency in what content would be blocked or removed:

> ‘I think in terms of restrictions for social media overall, I personally don’t feel they’re very open about exactly how they moderate posts. So I think that it’s difficult to understand what their thought processes [are] behind blocking posts or restricting posts because they don’t make it clear.’ (IDI)

Specifically, for SH/S content, participants emphasised that current platform regulations were too reactive, resulting in punitive measures such as immediate bans or content removal, where a more nuanced approach is needed to ensure support is provided to those who needed it:

> ‘…they seem to throw the baby out the bath water. They do a formal, catch you all sort of ban process there or suspend here…But where it comes to suicide and self-harm maybe it’s a point of that actually this is a serious situation where we need to be looking at this openly… rather than actually someone’s put themselves on the line to say ‘I’m feeling suicidal.’ All of a sudden it’s all guns blazing, red lights etcetera.Let me try and… have more safety-conscious rules and policies in place, that’s what we need to have.’ (IDG)

IDI echoed the call for a more thoughtful and empathetic approach to moderating online SH/S content, specifically highlighting the portrayal of SH scars. They emphasised a need to recognise scars can become a part of someone’s image and identity, and so removing the content would be disregarding their experiences and self-expression:

> ‘it’s like they’ve got a one size fits all rule thing where any posts about self-harm, any scars at all, they’ll just remove the pictures, whereas I’ve seen some people talk about how their scars are like recovery scars and just a part of who they are. So I think that improvement is still needed at the moment even just to establish the baseline of what’s right and what’s wrong to post about.’ (IDI)

A more multidimensional approach was encouraged for companies making decisions around SH/S content policy:

> ‘we need to get the social media firms to get their position right on what content they’re allowing to show on their platforms. They need to be real as well and actually work with local, national governments, but they also need to be working with like voluntary and community sectors of people that live with it. And also people with lived experience as well to actually find balance.’ – IDG

However, other participants favoured more rigorous moderation, particularly to protect vulnerable users, such as young people, who they acknowledged will seek out SH/S content and need safe spaces to engage:

> ‘If it was my child, I wouldn’t want them accessing that content but a lot of that is because I think that children and the internet are a difficult match anyway because of the way that the internet is designed to keep you hooked and put so much worth in algorithmic likes and comments.’ (IDF)

> ‘I feel like if teenagers or young people are gonna go on it [SH/S spaces online] then they need somewhere that will moderate it by professionals like I had when I were a teenager, rather than just being on Facebook or Instagram for example.’ (IDD)

One participant, who was exposed to graphic self-harm imagery on a mental health Facebook group noted that approving posts before they are shared could be an easily implemented moderation method:

> ‘…because on Facebook if you’re an admin of a group, because I do it on my group… you actually have to have admin approval before you approve it.’ (IDG)

> ‘Before other people see it’ (Interviewer)

> ‘Before it goes on the group. Now this [mental health Facebook] group doesn’t have that and this is the thing that was worrying’ (IDG)

##### 2.2.3.3. Professional vs. Lived Experience Moderators

Some participants went on to describe the moderator role in online spaces and whether this should be undertaken by trained professionals or by those with lived experience. Two of those participants expressed the importance of moderators with personal experience of SH/S:

> ‘That’s the great thing about [Self-Harm support Facebook group is] that it’s not run by people that don’t get it.’ (IDB)

> ‘I think having people have a lived experience doing the moderation is more beneficial.’ (IDC)

The general belief was that these moderators would have a better understanding of the content posted in the online communities allowing them to make informed judgements about the appropriateness of content, in context:

> ‘it’s not like you need like a psychiatrist or a counsellor on there or something like that. You know you don’t need that you just need somebody that’s been accountable for it… you know that’s not just some random person.’ (IDB)

Despite this, one individual preferred the concept of mental health professionals moderating, or for peer moderators to undergo training:

> ‘It definitely needs to have someone who’s trained in either like mental health first aid and specifically suicide and self-harm or like a proper professional’ (IDJ)

Participants also highlighted the importance of implementing mechanisms to prevent those with lived experience becoming too entrenched in a community’s mindset, which may be harmful to them. One suggestion was to have regular changes in moderators as a preventative measure. Another was to provide support to moderators from mental health professionals, to ensure they could handle content they may encounter, and maintain their own mental wellbeing:

> ‘I think there also needs to be somewhere to go to keep themselves in check because I think you can get just indoctrinated into what you’re doing, [with] everybody else on the site [having] the same problem. So whether it be something where moderators are changed regularly I don’t know, something like that.’ (IDH)

> ‘it feels like you need maybe somebody [to take] a step back. It may be that [Self-Harm Support Group] needs a professional that’s going to regularly contact the people that are moderating and the people that are running these things to make sure we’re okay.’ (IDB)

##### 2.2.3.4. Participants as Moderators

Three (IDA, IDB, IDI) had become moderators or administrators on the platforms where they accessed SH/S content. The decision to take on this role was driven by a hope to help others, and ensure rules within the online spaces were adhered to, as well as an altruistic way of giving back to the community:

> ‘If I can help support that even a little bit to make it a bit safer and keep people safe that are also vulnerable, it’s just my little way of saying thank you, I guess, for the support I’ve got and enabling other people to be able to still continue to get that support.’ (IDB)

These participants reflected on the significant responsibility associated with moderating online SH/S content. IDI shared their personal recognition of the need to disengage from a moderation/administrator role when their own mental health declined. They expressed concern about their ability to effectively address someone else’s issues in such circumstances, as well as the potential negative impact of engaging with another users’ distressing content:

> ‘I think the other thing is I won’t open a DM unless, again, I’m in a mood where I think I’ll be able to handle it because I don’t want a clash of negativity or anything to end up going wrong and, like you said, it’s the whole responsibility thing. So you don’t want someone to hear your point of view when you’re feeling negative yourself’ (IDI)

However, IDB seemed less able to manage their behaviour in this way. They recalled an incident where they encountered a video in the group they moderated that presented a SH method that was novel to them. This resulted in them subsequently acquiring the equipment for the method, and then self-harming:

> ‘Obviously, it [SH method post] got taken down from the main group… I’m first line of defence. It’s just that I keep getting hit by it at the moment because I’m the one that’s moderating it…it was talk about [SH method] and since then, I’ve bought [SH equipment] to try and do it myself.’ (IDB)

IDB, when considering the best approach to moderation, drew upon their own experience of encountering strict moderation methods. They found this experience influenced their gentler approach, recognising how more forceful moderation could potentially isolate vulnerable users, making them feel worse:

> ‘Different people do it different ways. I’m normally quite gentle because quite often, especially with the [Self-Harm Support Group] rules, the reason they’re breaking the rules is from a really nice place…We have got some people on the other end of the spectrum who say, ‘You agreed to the rules when you joined this group. You are breaking the rules. This is your one and only warning.’ I’ve been on the receiving end of that one and my first warning was like that and it made me nearly leave the group entirely…’ (IDB)

Additionally, IDI emphasised their approach to moderation involved being open and transparent, a quality they had criticised platforms for lacking in their own moderation practices:

> ‘I think if I thought that a comment would be damaging to someone else, then I’d delete it and then I’d message the person who sent the message to explain why I’ve done that.’ (IDI)

## 3. Discussion

Findings from this study suggest that although users engaging with SH and S content online favour the use of moderation and moderators in these spaces, the current methods used by platforms and communities may be inadequate to provide a safe and effective user experience due to inconsistencies, ambiguities, and biases

Many online environments rely on user reporting to ensure moderators can successfully enforce community rules [3]. However, participants in this study revealed an inherent inability to undertake community moderation through reporting SH/S content, particularly during times of poorer mental health. Participants revealed these struggles to report content stemmed from a fear of potentially stigmatising other users, and a desire to access the content themselves, despite recognising the potential dangers of the material. This emphasises the complexity of decision-making processes for vulnerable users in online spaces and challenges the traditional methods of content reporting strategies employed by platforms.

Although participants described difficulties in reporting SH/S content, several users did take on moderating roles in these online environments, with largely altruistic intentions. However, they also found their ability to moderate fluctuated alongside their own mental health, a concern recognised in previous research [6, 12]. While one user was able to identify their mental health declines and take action to protect themselves and others’ by temporarily withdrawing from their moderating/administrative duties, another was unable to safeguard themselves and became at risk of harm whilst undertaking their moderating role.

When considering SH/S content moderation by others, participants in this study described a lack of consistency in how platforms and community moderators responded to SH/S content. Additionally, poor transparency in communication of moderation methods and results not only led to disappointment, frustration, and a perceived loss of control amongst participants, but also added a speculative element to many participant accounts. Several also criticised ‘catch-all’ reactive policies for SH/S material, where content could be immediately removed, or users banned without warning. These punitive moderation methods were viewed as non-empathetic, harmful, and potentially stigmatising, consistent with user experiences of Facebook and Instagram’s ban of graphic self-harm imagery [5].

Participants largely agreed that lived-experience moderation was important, due to the moderators’ understanding of user experiences. However, many also emphasised the potential difficulties and harmful consequences a lived-experience moderator may experience by engaging with content and users in these spaces. Therefore, alike to participants in previous research [6] and recommendations from Facebook [16], some participants encouraged the involvement of mental health professionals in providing support through supervision or training, to lived-experience moderators.

### 3.1. Limitations

Using data from the DELVE research study (Haime et al., 2023) presented a valuable opportunity to explore user experiences of moderation, moderators, and moderating of SH/S content. This study is particularly relevant in the context of ongoing changes in legislature, such as the implementation of the Online Safety Bill [17] in UK law and the Digital Services Act [20] in Europe. Despite this, it is essential to acknowledge certain limitations in our approach. This study aimed to recruit participants who had engaged with SH/S content online. It was evident that participants encountered such content via several different platforms. Although this gave us good diversity, allowing for a broad understanding of user experiences, it made more nuanced platform-specific insights and implications difficult. Another limitation of this study was that participants who undertook moderator roles, were doing so voluntarily at the community-level and therefore we lacked insights from platform-level moderators, of moderating SH/S content. Additionally, where participants considered lived experience vs. mental health professional moderators in their accounts, they did not explicitly consider mental health professionals with lived experience of mental illness, potentially overlooking a factor affecting moderator effectiveness. Finally, we also struggled to recruit any participants who publicly posted the most explicitly harmful SH/S content (including graphic images/videos, and methods information) online. Exploring this perspective could offer valuable insights into participant perceptions regarding content reporting and removal, and how they respond to these actions.

### 3.2. Conclusions

Findings from this study should inform moderation practices by online industry leaders, with the following considerations:

- Platforms and communities should recognise the challenges faced by mental health online community members in practicing self-moderation. Reliance on user-reporting should be considered insufficient for providing a safe environment.
- Platforms and communities should consider integrating support from mental health professionals into mental health online spaces. Specifically, lived-experience moderators should have access to training and supervision from such professionals.
- Platforms and communities should consider the ongoing well-being of moderators. In doing so, they have a responsibility to ensure moderators have the capacity to recognise their own mental health concerns. This includes re-evaluating current models to ensure adequate time and mechanisms are allocated to moderators to address their mental and physical health needs.
- Platforms and communities should adopt a balanced approach to moderation of SH/S content, prioritising the safety of the overall userbase whilst also considering the wellbeing of individuals posting content. Platforms should allow for nuance in moderator decision-making processes, and prioritise the use of clear language and messaging to users around decisions made. They should also consider provision of postvention support to users following content removal or account banning.
- Platforms and communities should consider using an open dialogue approach with users in their moderation practices. This will enable users to stay informed of the processes their content or account may be undergoing and ensure transparency from platforms and communities regarding their moderation policies and guidelines.

These considerations may also provide insight to policymakers, such as Ofcom, in their role governing digital industry leaders. Policymakers should recognise the challenges online platforms and communities face in moderation of mental health spaces, and advocate for strategies that prioritise user safety. Policymakers should support the initiatives outlined above for industry leaders, promoting a transparent and sensitive approach to moderation of SH/S content online.

Findings from this study should also encourage further research into user experiences of moderation, moderators, and moderating in online mental health spaces. Exploring the perspectives of platform-level moderators overseeing SH/S content will further our understanding of moderation and moderators in these environments.

## 4. Method

### 4.1. Procedure

Data collected as part of the DELVE study [19] were analysed. Participants were interviewed at three time points (baseline, midpoint, endpoint) over a six-month study period about their engagement with SH/S content online. Interviews were semi-structured, with researchers employing an open-ended approach using probing to obtain detailed, participant-led accounts. In addition, participants were asked to maintain daily research diaries during the intervening periods. Participants were asked to reflect on their experiences of engaging with content, having content blocked or reported, or reporting others’ content during their interviews and in their diaries. In the interview, ‘moderation’ was an additional topic area where participants were asked to share their thoughts on moderators they had encountered online, their experiences of others’ content being moderated, and of their own SH/S content being moderated.

The study was approved by The University of Bristol Faculty of Health Sciences Ethics Committee (approval no. 117491). All participants provided written informed consent prior to participation.

### 4.2. Participants

Participants were 16 years and over, English-speaking, and online users who had engaged with self-harm or suicide content. Recruitment methods (outlined in [19]) involved multiple channels such as social media platforms, a mental health app targeting young people, and charity websites and newsletters. Participants responded to study adverts and were assessed for eligibility before being sent the study information sheet. Purposive sampling was used to promote diversity in gender, ethnicity, and age of sampled participants.

### 4.3. Analysis

Thematic analysis [18] with deductive coding informed by interview questions was used, with several steps:

1. Interviews were transcribed, and researchers read and familiarised themselves with both the participant transcripts and their diary data, to gain overall understanding.
2. ZH began coding the data. Codes were systematically assigned to quotes which captured certain concepts, or ideas about moderation or moderators. Coding was iterative, and codes were refined and revised in consultation with LB.
3. Similar codes were grouped together to form themes representing patterns in concepts or ideas. Themes were then explored narratively in the context of the research question.

## 5. Availability of Data and Materials

Anonymised transcript and questionnaire data will be stored on the University of Bristol’s Research Data Service Facility. Researchers will be able to request access to non-identifiable data upon reasonable request. Access will be subject to a data access agreement and following approval from Dr Lucy Biddle and the University of Bristol Data Access Committee.

## 6. Funding

This study was supported by the National Institute for Health and Care Research Bristol Biomedical Research Centre. The views expressed are those of the author(s) and not necessarily those of the NIHR or the Department of Health and Social Care. Original data collected as part of the DELVE study was funded by Samaritans, UK.

## 7. Competing Interests

We declare that there are no competing interests to disclose for any of the authors involved in the creation of this manuscript.

## 8. CRediT author statement

**Zoë Haime:** Project Administration, Investigation, Methodology, Formal Analysis, Writing (Original Draft), Writing (Review & Editing), Visualisation; **Lucy Biddle:** Funding Acquisition, Project Administration, Conceptualisation, Investigation, Methodology, Formal Analysis, Supervision, Writing (Review & Editing), Validation; **Laura Kennedy:** Investigation, Validation, Writing (Review & Editing); **Lydia Grace:** Conceptualisation, Writing (Review & Editing).

## 9. Acknowledgements

We express gratitude to the DELVE participants for their contribution to this work. Appreciation also extends to networks that facilitated recruitment, including TellMi, Samaritans, SMaRteN, McPin Foundation, and MQ Mental Health. We also acknowledge the contributions of our colleagues Jane Derges and Rachel Cohen, for their involvement in the DELVE study’s conception. We would also like to acknowledge our funders: National Institute for Health and Care Research Bristol Biomedical Research Centre and Samaritans, UK.

## Notes

### Competing Interest Statement

The authors have declared no competing interest.

### Funding Statement

Yes

